# Transactional sex and age-disparate sexual partnerships among adolescent girls and young women in Tanzania

**DOI:** 10.1101/2023.10.09.23296787

**Authors:** Katherine B. Rucinski, Gaspar Mbita, Kaitlyn Atkins, Esther Majani, Albert Komba, Caterina Casalini, Mary Drake, Neema Makyao, Anthony Galishi, Yeronimo Mlawa, Joyce Wamoyi, Sheree Schwartz, Stefan Baral, Kelly Curran

**Author notes:** Corresponding author: Katherine B. Rucinski 615 N. Wolfe Street, E5034 Baltimore, MD, USA.

## Abstract

Adolescent girls and young women (AGYW) continue to experience a high incidence of HIV in southern and eastern Africa, even in the context of large-scale HIV prevention interventions. In Tanzania, AGYW account for the largest proportion of new infections and have a higher risk of HIV acquisition than males of comparable age. We used routinely collected data from the PEPFAR/USAID-funded Sauti Project, a large combination HIV prevention program, to examine the relationship between transactional sex and sex with older partners among AGYW in Tanzania (2015-2020). Out-of-school AGYW 15-24 years completed a vulnerability index and were tested for HIV. We estimated weighted prevalence ratios (PR) and 95% confidence intervals (CI) for the associations of transactional sex (sex exchanged for money/services/gifts outside of sex work) and sex with older partners (≥5-years older, ≥10-years older) with prevalent HIV. We assessed potential synergism between exposures, and subgroup analyses explored associations among girls 15-19. 67,357 AGYW completed the vulnerability index and 14,873 had captured HIV testing records. Median age was 20 years (IQR 18-22). Transactional sex and age-disparate sex were common (35% and 28%, respectively); 13% of AGYW reported both behaviors. HIV prevalence was associated with both transactional sex (PR:1.28; 95% CI 1.00-1.63) and age-disparate sex (PR:1.26, 95% CI 0.99-1.60). In common referent analysis, transactional sex remained strongly associated with HIV, even in the absence of age-disparate sex (PR 1.41; 95% CI 1.02-1.94). Evidence of statistical synergism was not present, suggesting both transactional sex and age-disparate sex operate through similar pathways to increase HIV risk. Increased specificity within HIV prevention programs is needed to better meet the sexual and reproductive health needs of AGYW at high risk of HIV in Tanzania, including investment in tailored youth-friendly strategies for AGYW who have been marginalized from the current HIV response.

## Introduction

Over the last decade, substantial global investment and the expansion of critical prevention and treatment programs have resulted in overall declines in HIV incidence in countries across sub-Saharan Africa, including in Tanzania (1,2). Despite this progress, the burden of HIV remains high among key and other vulnerable populations such as adolescent girls and young women (AGYW) ages 15–24 (3). In 2022, AGYW account for the highest proportion of new infections in Tanzania and remain at greater risk of infection relative to their male counterparts and other age groups (1,4,5).

Structural, economic and social dynamics in eastern and southern Africa, along with increased biological vulnerability, heighten young women’s risk of acquiring HIV (6,7). During adolescence and early adulthood, key transitions including physical, physiological, emotional, and social changes happen rapidly, affecting relationship formation and sexual decision making (8–11). As young people navigate these changes, gender norms that assign greater social and economic power to men create the material and ideological conditions that make it difficult for young women to refuse sex, negotiate condom use, or advocate for their own sexual interests (12–15). These dynamics may be heightened for young women living in poverty, including AGYW that live in food-insecure households and are unable to readily meet their basic needs from sources other than boyfriends or other sexual partners (16).

Sexual relationships with older male partners can increase the risk of HIV for AGYW in eastern and southern Africa. These relationships, which often comprise a sexual relationship with a male partner ten or more years older, can also lead to transmission of other sexually transmitted infections, early pregnancy, and child marriage (17,18). In South Africa, increasing differences in partner age are associated with risk of HIV infection, particularly for adolescents (19). Phylogenetic studies also indicate that men 25–40 years are the main source of HIV transmission for AGYW ages 15–25 (20). Similar findings have emerged in Kenya, Uganda, and Zimbabwe, with partner age differences as little as five years predictive of elevated HIV risk (21,22).

Transactional sex, defined by the exchange of sex for money or material support, has also been shown to be consistently associated with HIV acquisition (23–25). Transactional sex exists across a continuum for AGYW, with exchanges occurring within established partnerships, casual encounters, or within the context of more formalized sexual exchanges such as sex work (26,27). AGYW who report transactional sex have also reported a high prevalence of emotional and sexual violence, substance use, and condomless sex. When transactional sexual relationships occur with older males, these HIV risks can be further amplified given a higher prevalence of infection (28–30).

Study of transactional sex with older partners among AGYW in eastern and southern Africa has increased over the last five years, with inferences largely made using established research cohorts in countries such as South Africa, Uganda, and Kenya (23,31,32). While substantial qualitative work has been done to contextualize experiences of transactional sex in Tanzania, few empiric studies have quantitatively examined the relationship between transactional sex and HIV in the context of age-disparate relationships (22,27,33,34). Here, we use routinely collected data from a large combination HIV prevention program to better define the relationship between transactional sex and sex with older partners, and their joint impact on the HIV burden among AGYW in Tanzania.

## Methods

### Study setting and program

Data included in this analysis are from the Sauti Project, a large community-based HIV prevention program funded by the United States Agency for International Development (USAID) through the President’s Emergency Plan for AIDS Relief (PEPFAR). Implemented by Jhpiego in partnership with EngenderHealth, Pact, and the Tanzania National Institute for Medical Research, Sauti provided a core package of client-centered sexual and reproductive health services for key populations such as female sex workers and men who have sex with men (35–39). Sauti also delivered critical HIV-prevention services for vulnerable AGYW ages 15-24 within 14 regions of Tanzania between 2015 and 2020, and was also an implementing partner of the DREAMS (Determined, Resilient, Empowered, AIDS-free, Mentored, and Safe) Initiative (**Figure 1**) (34,40). Programming delivered through Sauti for AGYW included biomedical services such as HIV testing and counseling, HIV case management, sexual risk assessments, family planning counseling and provision of contraception, screening for sexually transmitted infections (STI) and tuberculosis, nutritional assessments, and additional screening and referrals for substance use and gender-based violence. Further programming included behavioral change sessions and social protection interventions such as savings and loans clubs.

**Figure 1.**
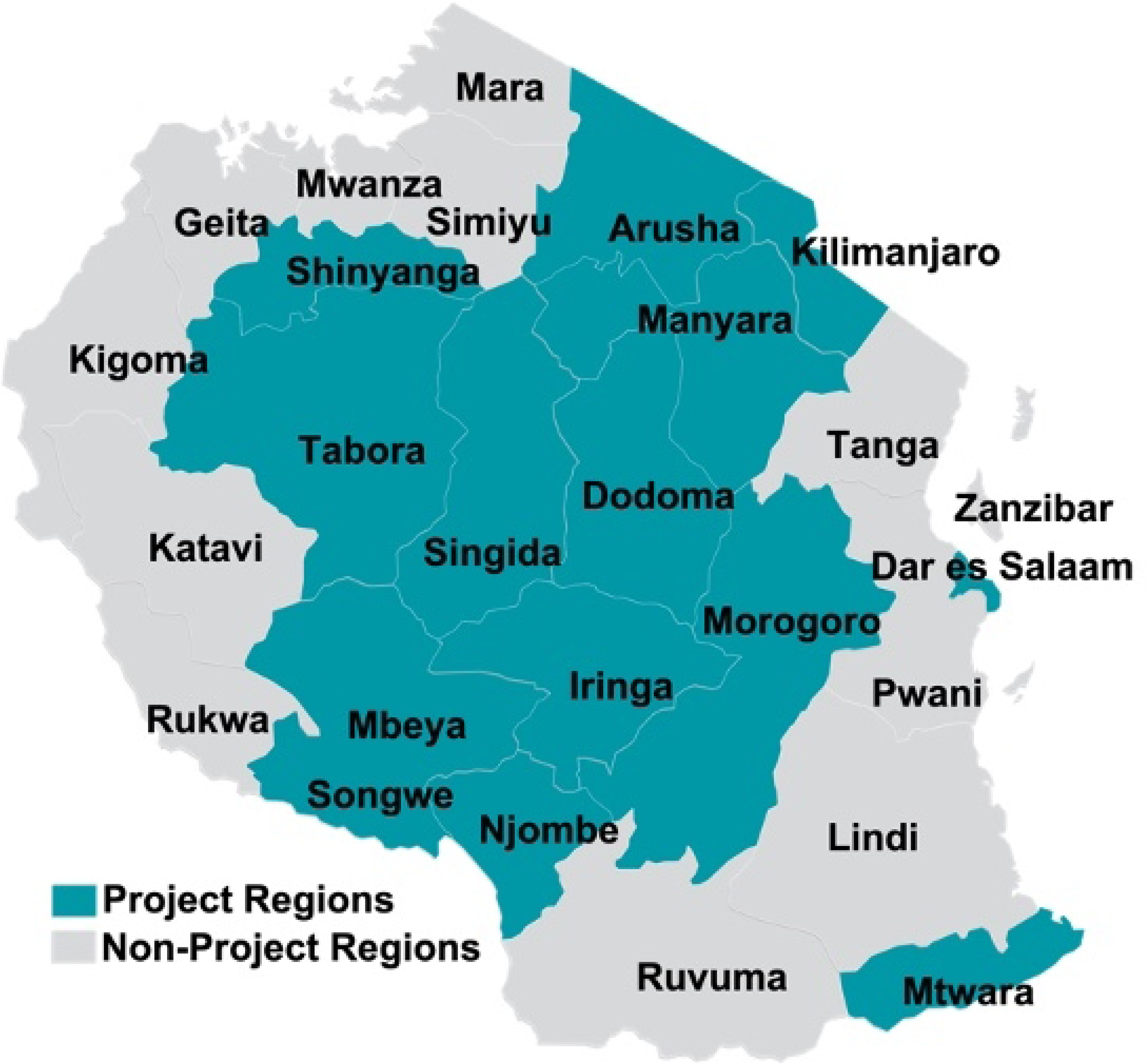
Sauti Project regions in Tanzania, 2015-2020

### Recruitment, eligibility, and procedures

AGYW between the ages of 15 and 19 who were not in school full time (defined as not having attended classes more than 10 days in the last three months when not on public holidays or school vacation) and those aged 20-24 were recruited beginning in 2015. Venues included biomedical service-delivery points such as community-based HIV testing and counseling sites, workplaces, bars, guest houses, salons, markets, other venues previously identified as “hotspots” by the program, or AGYW-identified DREAMS safe spaces including religious houses and local government offices. Trained peer facilitators recruited AGYW from each site and assessed their eligibility for program services using a brief tool. This vulnerability index was adapted using existing tools from Tanzania and other countries in eastern and southern Africa (40). Methods to develop this index have been previously described (40). Briefly, measures included basic demographics, indicators of sexual experience including partner number and age, engagement in transactional sex, experiences of intimate partner violence, depression, and food insecurity, among others. The full index is reported in the Supplemental Materials **(S1 File)**. Further, AGYW who accessed HIV testing and other biomedical services through Sauti also completed a health screening and service tool (HSST). Typically, these HIV testing visits occurred after AGYW had engaged in behavior change sessions, economic empowerment sessions or other Sauti biomedical services. Information routinely collected through the HSST included basic demographics, screening for harmful drinking of alcohol and substance use, STI screening and treatment, and each AGYW’s HIV testing results.

### Study population

We assembled an analytic sample using data collected among AGYW through the vulnerability index and merged these data with biomedical HIV testing records collected through the HSST using participant-specific alphanumeric codes. Data were restricted to those AGYW that accessed services through Sauti between 2015-2019, and to those AGYW that were eligible for inclusion based on the established eligibility criteria as described. Notably, some AGYW who did not meet full eligibility requirements, including those that were still in school, were also referred for Sauti services if they expressed interest in engaging with the program; these AGYW were excluded from analyses. In some cases, AGYW had duplicate or repeat vulnerability index records. For these AGYW, we used the earliest record available in the database. Similarly, some AGYW had multiple HIV testing results reported over multiple years. For these AGYW, we used the date of first HIV testing record under the assumption that this visit occurred closet to initial program engagement. Young women ages 18-24 who identified as female sex workers were also excluded from analyses, as they were ineligible to complete the AGYW vulnerability index.

### Outcome and Exposures

The primary outcome was prevalent HIV infection (yes/no), defined as a positive HIV test result documented through the Sauti HSST. Standard HIV testing comprised serial testing using the SD Bioline HIV-1/2 3.0 (Standard Diagnostics Inc., Suwon, Korea) and Uni-Gold HIV-1/2 (Trinity Biotech, Wicklow, Ireland) assays.

The two primary exposures included in this analysis were self-reported engagement in transactional sex and sex with older partners, both assessed through the vulnerability index. Transactional sex was defined as having ever engaged in any sex for money, services, or gifts (yes/no) (41). Age-disparate sex was defined as having engaged in sex with a partner five or more years older; intergenerational sex was defined as having ever engaged in sex with a partner ten or more years older (yes/no) (42,43).

### Statistical Analysis

Demographics and characteristics of AGYW participants who completed the vulnerability index were summarized using proportions for categorical variables and medians for continuous variables. Chi-square tests were used to compare the differences in proportions between groups for categorical variables, and Wilcoxon signed-rank sum tests were used for continuous data (alpha= 0.05).

We assessed differences in sample characteristics among participants who did and did not have a linked HIV testing result confirmed through the HSST. Among participants with an HIV testing result, we further assessed differences in history of transactional sex, comparing those who had engaged in transactional sex with those who reported never having engaged in transactional sex. We used a modified Poisson regression model with a robust variance estimator to estimate prevalence ratios (PR) and 95% confidence intervals (CI) for the association between transactional sex and prevalent HIV infection. Models were restricted to AGYW who were sexually active. A directed acyclic graph (DAG) was used to identify a minimally sufficient set of confounders for adjustment. Covariates in the minimally sufficient set included age, age-disparate sex/intergenerational sex, adult support, marital status, prior pregnancy, food insecurity, year, and early sexual debut. Models were similarly developed to assess associations between age-disparate sex and HIV, as well as intergenerational sex and HIV. Interaction was assessed between transactional sex and age-disparate sex, and between transactional sex and intergenerational sex using common referent analysis and through comparison of observed and expected joint effects (44). Subgroup analyses assessed main associations among adolescent girls ages 15-19.

### Weights

Given missing HIV testing outcomes among those AGYW that did not have a linked record through the HSST, we derived non-response weights under the assumption that these data were missing at random (MAR) to address the possibility of selection bias and to improve the internal validity of our findings (45,46). Weights were created using the full database of 67,357 unique and eligible AGYW participants who completed the vulnerability index. An indicator variable for having an observed versus a missing HIV testing result was created and estimated as a function of a participant’s measured covariates including age, partner status, prior pregnancy, food insecurity, sexual debut, experiences of violence, region, and survey version (year). Covariates were chosen based on preliminary analyses to identify patterns of missing information in these data, and we used multiple imputation by chained equations (MICE) to fill in missing data for all covariates (n=10 imputations) (47,48). We calculated and applied weights to all effect estimates for each series of imputations. Weights were stabilized by the marginal probability of having an observed HIV testing outcome and were truncated at the 5^th^ and 95^th^ percentiles to further improve stability. Weighted effect estimates and standard errors were pooled to calculate final PRs and 95% CIs, standardized to the full population of AGYW who completed the vulnerability index.

All analyses were conducted using SAS 9.4 (SAS Institute, Cary, NC).

### Ethics

The National Institute for Medical Research and the Ministry of Health Community Development, Gender, Elderly and Children of the United Republic of Tanzania provided ethical clearance for primary data collection activities under Sauti. The vulnerability index was administered in the context of routine service delivery, and all participants provided verbal informed consent. Parental consent was only sought for HIV testing if the participant was under the age of 18 in accordance with Tanzania law. Minors under the age of 18 who were parents were considered emancipated, and thus did not require parental consent. Ethical approval for the use of de-identified routine data was provided by the National Institute for Medical Research (NIMR/HQ/R.8c/Vol.1/678) and the Johns Hopkins Institutional Review Board (IRB No. 00006673).

## RESULTS

### Sample characteristics

A total of 67,357 unique AGYW participants completed the Sauti vulnerability index between 2015-2019 (**Figure 2**). Based on their age and schooling status, 65,185 were eligible for inclusion and retained in further analyses. Of these AGYW participants, 19,748 (30.4%) lived in Dar es Salaam, 11,240 (17.3%) in Mbeya, and 8,891 (13.7%) in Shinyanga; the remaining participants were from Arusha, Dodoma, Iringa, Kilimanjaro, Morogoro, Njombe, Songwe, and Tabora.

**Figure 2.**
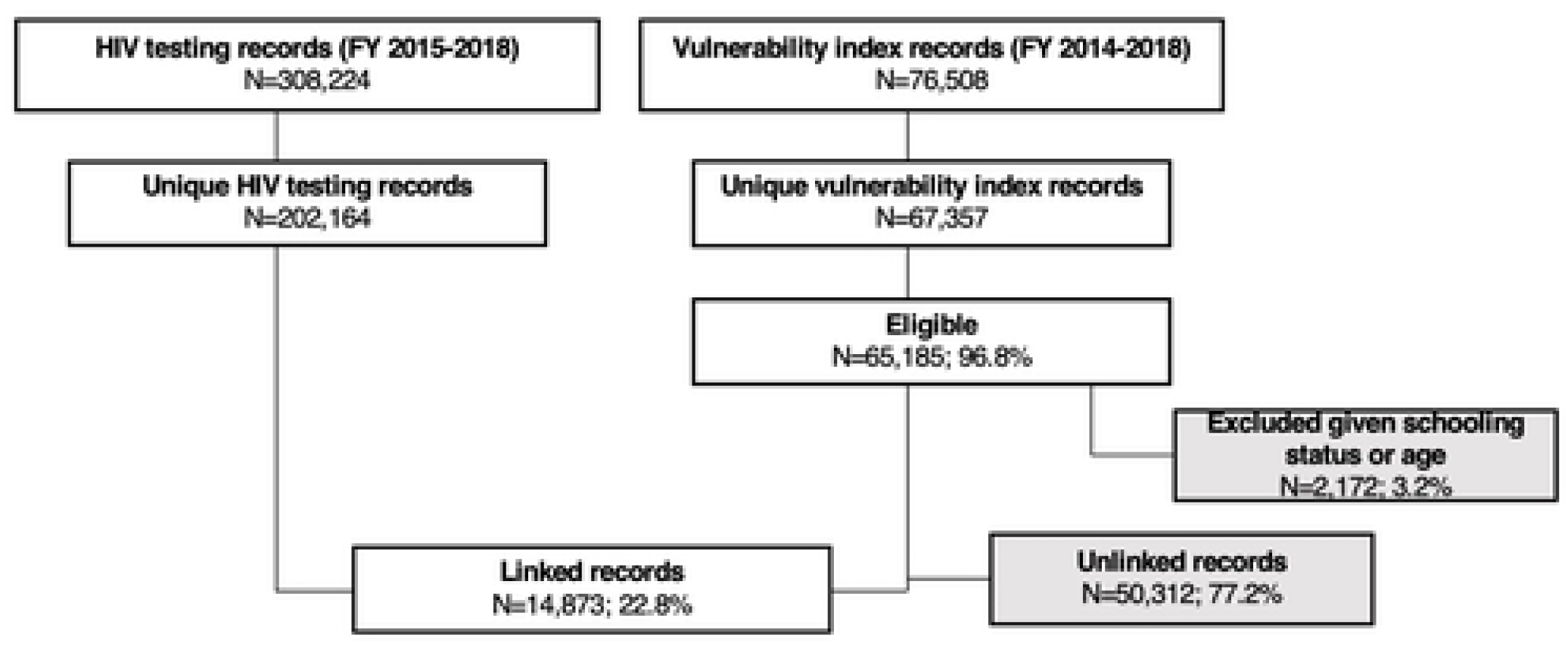
Construction of analytic sample including 14,873 AGYW using program data from the Sauti Project, 2015-2019

A total of 14,873 (22.8%) AGYW were able to be linked to their HIV testing record through the HSST using a unique identifier. Of these 14,873 AGYW, the median age was 20 years (IQR 15, 24) (**Table 1**). One-fifth (20.9%) were married. Almost half (40.7%) had previously been pregnant. Lifetime experience of transactional sex and age-disparate sex were common (35.6% and 39.1%, respectively). A total of 278 (1.9%) AGYW were living with HIV. Characteristics differed significantly by experience of transactional sex.

**Table 1.** Characteristics of 14,873 AGYW accessing combination HIV prevention services in Tanzania through the Sauti Project, 2015-2019^a^.

|  | Overall<br>N=14,873 |  | No transactional<br>sex<br>N=9,521 (65.4%) |  | Transactional sex<br>N=5,047 (34.6%) |  |  |
| --- | --- | --- | --- | --- | --- | --- | --- |
|  | Median | IQR | Median | IQR | Median | IQR | p-<br>value |
| Age, years | 20.0 | 18.0-22.0 | 20.0 | 18.0-22.0 | 21.0 | 19.0-23.0 | <.001 |
|  | n | % | n | % | n | % | p-<br>value |
| Married | 3,063 | 20.9 | 1,916 | 20.5 | 1,096 | 22.0 | <.001 |
| Adult support<sup>b</sup> | 7,949 | 55.0 | 4,381 | 47.1 | 3,482 | 70.2 | <.001 |
| Food insecurity<sup>c</sup> | 4,302 | 29.0 | 2,081 | 22.3 | 2,179 | 44.4 | <.001 |
| Ever pregnant | 6,035 | 40.7 | 3,369 | 35.4 | 2,587 | 51.3 | <.001 |
| Sexual debut <15 years | 2,544 | 17.7 | 1,099 | 11.8 | 1,412 | 28.7 | <.001 |
| Sexual partner >5 years older | 4,162 | 28.3 | 2,156 | 22.8 | 1,953 | 39.1 | <.001 |
| Sexually active, ever | 12,730 | 86.4 | 7,509 | 79.0 | 5,006 | 99.9 | <.001 |
| Sexually active, last 12 months | 10,895 | 76.1 | 6,230 | 67.0 | 4,522 | 92.9 | <.001 |
| Using modern contraception<sup>d</sup> | 4,774 | 36.8 | 2,663 | 31.5 | 2,111 | 46.5 | <.001 |
| Condomless sex, last 12<br>months<sup>e</sup> | 791 | 74.1 | 472 | 68.0 | 319 | 85.5 | <.001 |
| >1 partner, last 12 months | 5,014 | 34.8 | 1,777 | 19.0 | 3,127 | 65.4 | <.001 |
| Sexual violence, ever | 1,901 | 13.1 | 771 | 8.2 | 1,109 | 22.4 | <.001 |
| Sex with HIV+ partner, last 12<br>months | 411 | 2.9 | 186 | 2.0 | 216 | 4.4 | <.001 |
| Prevalent HIV | 278 | 1.9 | 150 | 1.6 | 125 | 2.5 | .006 |

<sup>a</sup> Missing. transactional sex: 305 (2.1%); married: 244 (1.6%); adult support: 427 (2.9%); food insecurity: 484 (3.3%); pregnancy: 25 (0.2%); sexual debut: 470 (3.2%); partner age: 153 (1.0%); ever sexually active: 547 (3.7%); sexually active in the last year: 139 (0.9%); contraception: 659 (4.8%); number of sex partners: 438 (2.9%); sexual violence: 390 (2.6%); sex with HIV+ partner: 438 (2.9%); HIV: 341 (2.3%)
<sup>b</sup> Has an adult in the household or community who provides unconditional emotional and/ or financial support
<sup>c</sup> Could not afford to buy food or there was not enough food to eat at home
<sup>d</sup> Use of modern contraception asked on versions 1 and 2 of the index
<sup>e</sup> Used a condom never, almost never, or sometimes when having vaginal sex; asked on version 3 of the index

The remaining 50,312 participants did not have a matching HIV testing record in the database. Compared to those for whom an HIV testing record was not available, AGYW with observed HIV testing outcomes were less likely to be married (20.9% vs. 23.6%; p<0.001), to have experienced recent food insecurity (29.9% vs. 40.8%; p<0.001), to have ever experienced sexual violence (13.1% vs. 21.8%; p<0.001), and to have engaged in transactional sex (37.0% vs. 45.1% p<0.001). Characteristics among AGYW participants with and without a linked HIV testing record are reported in **S1 Table**.

### Transactional sex and HIV

Lifetime experience of transactional sex was associated with HIV prevalence in unadjusted (PR: 1.41; 95% CI 1.10, 1.80) and adjusted models (PR: 1.34; 95% CI 1.04, 1.74) (**Table 2**). Weighted estimates similarly demonstrated a population-level association between transactional sex and HIV (PR: 1.28; 95% CI 1.00, 1.63).

**Table 2.**
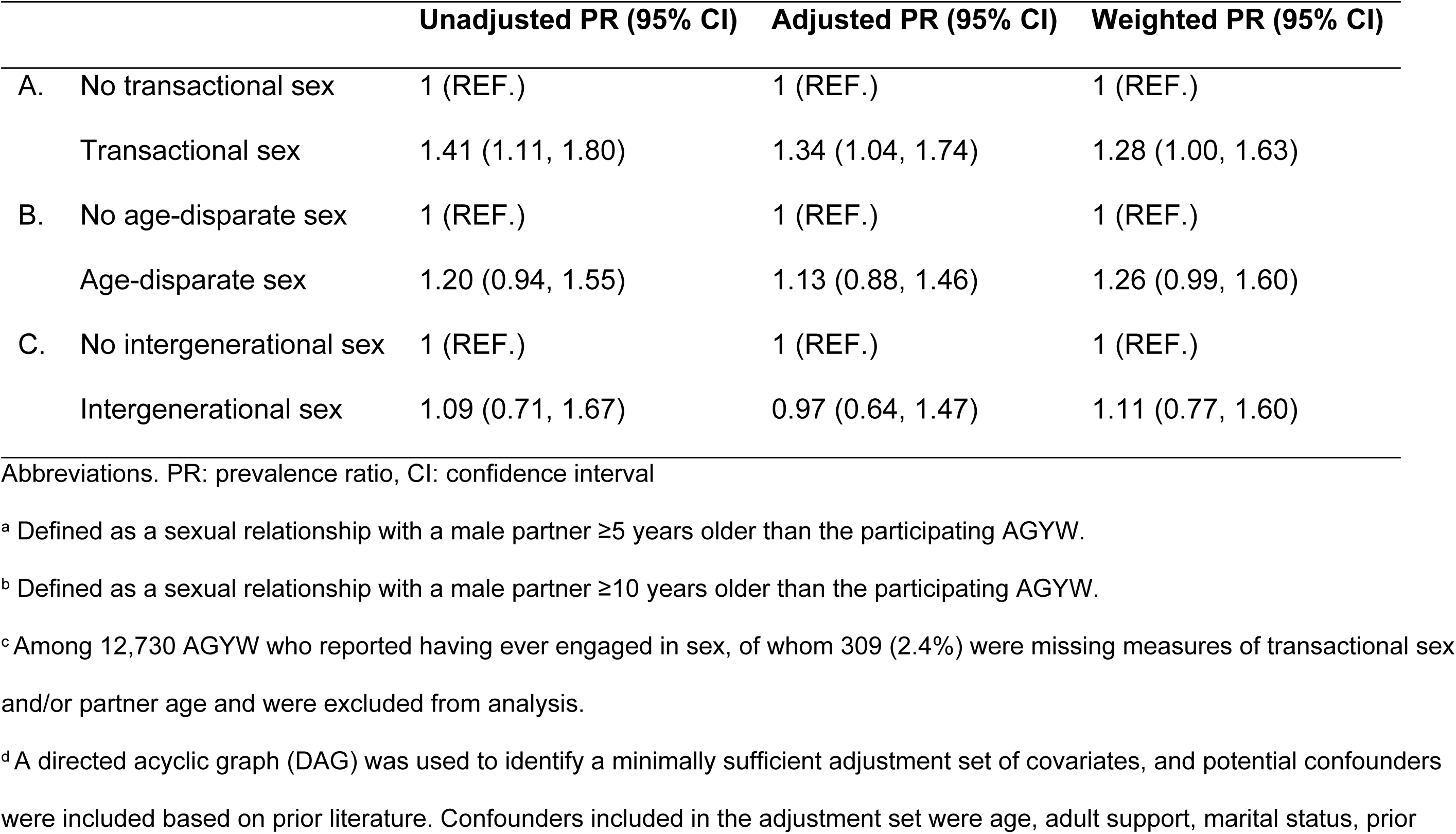
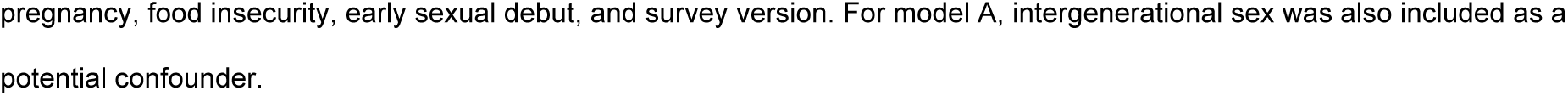
Unadjusted, adjusted, and weighted associations of A) transactional sex with HIV prevalence; B) age-disparate sex^a^ with HIV prevalence; and C) intergenerational sex^b^ with HIV prevalence among 12,730 sexually active AGYW accessing combination HIV services in Tanzania through the Sauti Project, 2015-2019^c,d^.

|  | Unadjusted PR (95% CI) | Adjusted PR (95% CI) | Weighted PR (95% CI) |
| --- | --- | --- | --- |
| A. No transactional sex | 1 (REF.) | 1 (REF.) | 1 (REF.) |
| Transactional sex | 1.41 (1.11, 1.80) | 1.34 (1.04, 1.74) | 1.28 (1.00, 1.63) |
| B. No age-disparate sex | 1 (REF.) | 1 (REF.) | 1 (REF.) |
| Age-disparate sex | 1.20 (0.94, 1.55) | 1.13 (0.88, 1.46) | 1.26 (0.99, 1.60) |
| C. No intergenerational sex | 1 (REF.) | 1 (REF.) | 1 (REF.) |
| Intergenerational sex | 1.09 (0.71, 1.67) | 0.97 (0.64, 1.47) | 1.11 (0.77, 1.60) |

Abbreviations. PR: prevalence ratio, CI: confidence interval
<sup>a</sup> Defined as a sexual relationship with a male partner $\geq 5$ years older than the participating AGYW.
<sup>b</sup> Defined as a sexual relationship with a male partner $\geq 10$ years older than the participating AGYW.
<sup>c</sup> Among 12,730 AGYW who reported having ever engaged in sex, of whom 309 (2.4%) were missing measures of transactional sex and/or partner age and were excluded from analysis.
<sup>d</sup> A directed acyclic graph (DAG) was used to identify a minimally sufficient adjustment set of covariates, and potential confounders were included based on prior literature. Confounders included in the adjustment set were age, adult support, marital status, prior
pregnancy, food insecurity, early sexual debut, and survey version. For model A, intergenerational sex was also included as a potential confounder.

### Older partners and HIV

There was a moderate association between age-disparate sex and HIV in unadjusted analysis (PR: 1.20; 95% CI 0.94, 1.55) that was attenuated when adjusted for potential confounders (PR: 1.15; 95% CI 0.89, 1.50). In weighted analysis, age-disparate sex was associated with a 26% increase in HIV prevalence (PR: 1.26; 95% CI 0.99, 1.60). Intergenerational sex was not strongly associated with HIV prevalence in unadjusted (PR: 1.09; 95% CI 0.71, 1.67), adjusted (PR: 1.00; 95% CI 0.65, 1.53), or weighted models (PR: 1.11, 95% CI 0.77, 1.60).

### Combined association of transactional sex and sex with older partners

In common referent analysis, AGYW who engaged in transactional sex in the absence of age-disparate partners had 1.41 (95% CI 1.02, 1.94) times the prevalence of HIV compared to AGYW who reported neither behavior (**Table 3**). Conversely, age-disparate sex was not associated with HIV prevalence in the absence of transactional sex (PR: 1.19, 95% CI 0.81, 1.67). The prevalence ratio for those who reported both transactional sex and age-disparate sex was 1.45 (95% CI 1.01, 2.08), which was less than the expected prevalence ratio using both additive and multiplicative criteria.

**Table 3.**
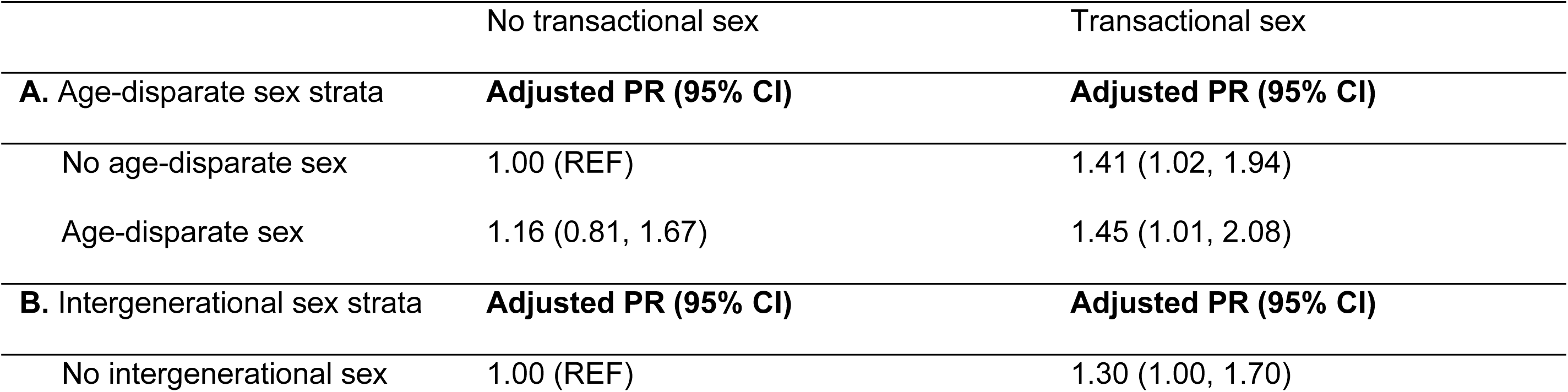
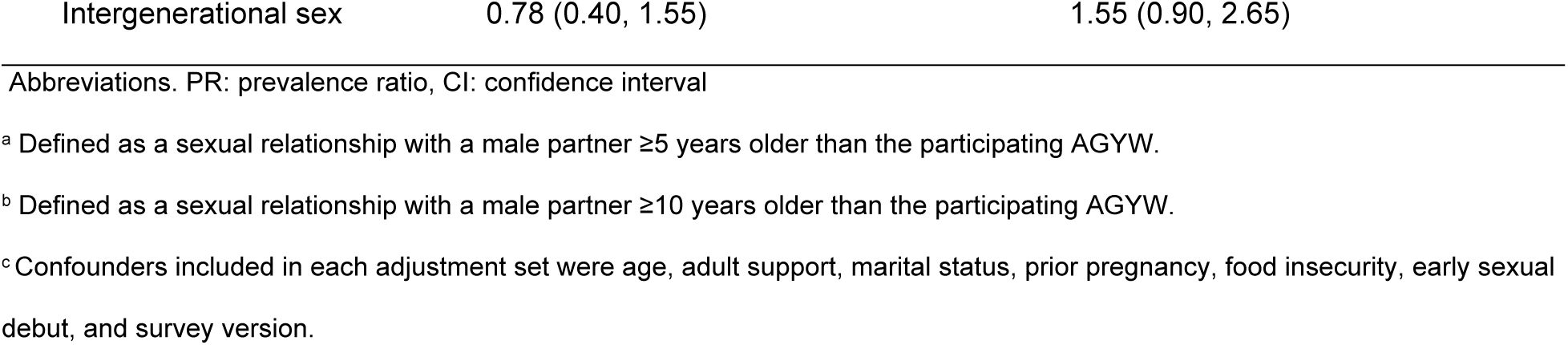
Interaction between a) transactional sex and age-disparate sex^a^ and b) transactional sex and intergenerational sex^b^ on HIV prevalence among 12,730 sexually active AGYW in Tanzania, 2015-2019^c^.

### Subgroup analysis among AGYW age 15-19

Transactional sex was also common among AGYW age ≤19 (**Table 4**). Transactional sex, age-disparate sex, and intergenerational sex were not significantly associated with HIV prevalence among younger AGYW (**S2 Table**).

**Table 4.** Characteristics of 5,816 adolescent girls ages 15-19 accessing combination HIV prevention services in Tanzania through the Sauti Project, 2015-2019^a^.

|  | Overall<br>N=5,816 |  | No transactional<br>sex<br>N=3,879 (68.2%) |  | Transactional sex<br>N=1,813 (31.9%) |  |  |
| --- | --- | --- | --- | --- | --- | --- | --- |
|  | Median | IQR | Median | IQR | Median | IQR | p-<br>value |
| Age, years | 18.0 | 17.0-19.0 | 18.0 | 17.0-19.0 | 18.0 | 17.0-19.0 | <.001 |
|  | n | % | n | % | n | % | p-<br>value |
| Married | 553 | 9.7 | 320 | 8.4 | 219 | 12.3 | <.001 |
| Adult support<sup>b</sup> | 3,015 | 53.5 | 1,758 | 46.5 | 1,230 | 68.9 | <.001 |
| Food insecurity<sup>c</sup> | 1,735 | 30.9 | 899 | 23.8 | 820 | 46.2 | <.001 |
| Ever pregnant | 1,321 | 22.7 | 679 | 17.5 | 627 | 34.6 | <.001 |
| Sexual debut <15 years | 1,310 | 23.2 | 594 | 15.6 | 702 | 39.5 | <.001 |
| Sexual partner >5 years older | 1,282 | 22.3 | 648 | 16.8 | 617 | 34.4 | <.001 |
| Sexually active, ever | 4,272 | 74.3 | 2,399 | 61.9 | 1,800 | 100.0 | <.001 |
| Sexually active, last 12 months | 3,576 | 63.7 | 1,913 | 50.3 | 1,618 | 92.1 | <.001 |
| Using modern contraception<sup>d</sup> | 1,363 | 26.8 | 706 | 20.6 | 657 | 39.6 | <.001 |
| Condomless sex, last 12 months<sup>e</sup> | 222 | 70.3 | 136 | 63.3 | 86 | 85.1 | <.001 |
| >1 partner, last 12 months | 1,657 | 29.4 | 577 | 14.1 | 1,074 | 60.9 | <.001 |
| >2 partners, last 12 months | 526 | 9.3 | 162 | 4.2 | 363 | 20.6 | <.001 |
| Sexual violence, ever | 673 | 11.9 | 307 | 8.1 | 355 | 19.9 | <.001 |
| Sex with HIV+ partner, last 12 months | 163 | 2.9 | 76 | 2.0 | 8 | 4.6 | <.001 |
| Prevalent HIV | 65 | 1.2 | 34 | 0.9 | 29 | 1.7 | .002 |

<sup>a</sup> Missing. Married: 103 (1.8%); adult support: 185 (3.2%); food insecurity: 202 (3.5%); pregnancy: 5 (0.1%); sexual debut: 180 (3.1%); partner age: 58 (1.0%); ever sexually active: 68 (1.2%); sexually active in the last year 204 (3.5%); number of sex partners: 174 (3.0%); sexual violence: 176 (3.0%); sex with HIV+ partner: 165 (2.8%); HIV: 191 (3.3%)
<sup>b</sup> Has an adult in the household or community who provides unconditional emotional and/ or financial support
<sup>c</sup> Could not afford to buy food or there was not enough food to eat at home
<sup>c</sup> Could not afford to buy food or there was not enough food to eat at home
<sup>d</sup> Use of modern contraception asked on versions 1 and 2 of the index
<sup>e</sup> Used a condom never, almost never, or sometimes when having vaginal sex; asked on version 3 of the index

## DISCUSSION

In this study, we examined the relationship between HIV burden and transactional sex among over 14,000 AGYW accessing real-world HIV prevention services through the PEPFAR/USAID-funded Sauti Project in Tanzania. Transactional exchanges of money, services, or gifts for sex were common among AGYW enrolled in Sauti programing, including among girls as young as age 15. These exchanges occurred among AGYW who were mostly unmarried, had previously been pregnant, and were receiving emotional and financial support from older adults, including family and other community members. AGYW who reported transactional sex had an elevated burden of HIV, and transactional sex was strongly associated with HIV prevalence even in the absence of older partners. Given the multiple social and structural vulnerabilities that can lead to transactional sexual relationships, including those with older partners, there is need to identify HIV prevention approaches that are more responsive to AGYW who may necessitate tailored and targeted programmatic support early in adolescence.

The burden of HIV was not evenly distributed among AGYW in this analysis, with estimates of HIV prevalence the highest among young women who reported both transactional sex and sexual relationships with older partners. While intergenerational and transactional sex are often conceptualized in the literature as compatible risk factors for HIV acquisition (23,24,29,49), few studies have explored the extent to which asymmetry in partner age and transactional sex interact to increase HIV-associated vulnerability (28). Consistent with our findings, one case-control study in South Africa found women who reported both transactional sex and intergenerational partners had nearly twice the odds of HIV compared to women who reported either experience alone (22). These findings contribute to a growing body of literature that aim to further distinguish heterogeneities of risk among AGYW and more effectively guide HIV prevention programming. Importantly, early findings from the DREAMS initiative have demonstrated little to no improvement in reducing HIV incidence across multiple countries in southern and eastern Africa (50). Our results suggest that aligning HIV prevention services with the needs of AGYW who necessitate more targeted and intense approaches-such as young women who engage in resource-driven transactional relationships with older partners-are needed to deliver meaningful reductions in incidence for AGYW moving forward. Approaches may include economic empowerment for broader social asset building (51), integrated family planning and maternal health services to support the needs of adolescent mothers (52), as well as vocational training to facilitate employment and improve income (53), among others.

Experiences of transactional sex are often predicated on structural vulnerabilities, gender norms, and power differentials that motivate AGYW to engage in resource-driven sexual relationships. The high burden of food insecurity, prior pregnancy, and sexual violence among AGYW in this study, particularly among those reporting transactional sex, reinforce the larger social and structural determinants of HIV that underly sexual exchanges and increase risk of infection. For AGYW living in poverty, transactional sex offers a means to provide food, clothing, shelter and other resources, and can establish pathways to support and security. These more explicit transactional relationships may be even riskier for younger AGYW, who may be further marginalized or restricted from accessing sexual and reproductive health services due to their age (54). While transactional sex and sex with older partners were not strongly associated with HIV prevalence among adolescents in subgroup analyses, an alarmingly high prevalence of condomless sex among girls as young as 15 engaged in transactional sex indicates a potential high incidence of infection in the context of these larger social and structural factors (7,49,55).

AGYW are broadly considered a priority population for HIV epidemic control, but not all AGYW have a high risk of acquiring HIV. While young women >18 years who identified as female sex workers were explicitly excluded from analyses, a high prevalence of condomless sex, violence, and food insecurity among adolescent girls accessing Sauti services suggests some overlap with sex work. Notably, few programs in southern and eastern Africa have evaluated the impact of tailored HIV programming for young women who sell sex (56–58). In Zimbabwe, DREAMS programming that implemented community mobilization and social protection interventions alongside dedicated HIV testing and PrEP for young women who sell sex found increased uptake of clinical services; however, no effect on HIV incidence was observed (59). Additional efforts in Zimbabwe, Mozambique, and Kenya have worked to engage young women who sell sex, but more broadly this population remains under-represented in both research and programs (58,60). This gap may be further perpetuated by issues of identity. Young women who sell sex may not identify as sex workers until they are older (61,62), but may be marginalized from existing AGYW HIV prevention and treatment programs (16,63,64). They may also feel ostracized by older sex workers, thus limiting the extent to which they engage in FSW HIV prevention and treatment efforts such as drop-in centers. Thus, there is an urgent need to engage this population in both research and program design efforts, and to grow the evidence base of effective interventions for young women who engage in necessity-driven transactional sex.

A strength of this study was our ability to leverage a large database of observational data collected during the delivery of routine services through the Sauti Project in Tanzania. In general, population-level inferences made using programmatic data tend to be more representative than research data, which generally are collected using narrow eligibility criteria that may threaten external validity (65). Well-controlled research studies may also exclude adolescents who are most at risk for poor sexual and reproductive health outcomes such as those who live and work on the street, potentially limiting generalizability of findings (66,67). Yet programmatic data remain a relatively under-utilized resource to examine the effectiveness of HIV-related interventions and implementation strategies for key populations (68–71). As programs continue to roll out biomedical prevention interventions for AGYW such as long-acting PrEP, program data offer an opportunity to keep pace and quickly tailor and adapt implementation strategies, thus providing for a more nuanced and more efficient HIV response.

This study also had several limitations. Our use of program data necessitated the merging of multiple databases. Rigorous quality assurance procedures were implemented throughout the duration of the program; however, challenges with unique identifiers limited the number of AGYW who could be linked across program records. We used non-response weights to try and improve internal validity under the assumption these unlinked records were MAR given the observed patterns in which the “missingess” of records occurred. Additionally, changes to the vulnerability index and question format during program implementation precluded us from fully assessing key measures including frequency of condom use, which may have provided additional insight into behavioral risks within transactional sexual partnerships. Further, we were also limited by temporality and were unable to determine if transactional or intergenerational sexual partnerships were more recent, and whether HIV was sexually or pediatrically acquired. Nevertheless, these findings provide critical insights into multiple relationship dynamics among AGYW in Tanzania. Given relatively few research data available for AGYW in the context of HIV prevention and treatment uptake in Tanzania (72), these estimates fill an epidemiologic gap and can inform targets for intervention moving forward.

## Conclusions

Declines in HIV incidence among AGYW over the last several years have been smaller than expected given substantial investments in HIV programming for AGYW in eastern and southern Africa through large combination HIV prevention programs such as DREAMS. This study contributes to a growing body of evidence that HIV risks among AGYW are not homogenous, and that additional implementation strategies are needed to link AGYW at high-risk of HIV to behavioral, structural and biomedical prevention interventions such as PrEP. These efforts likely include increased specificity among AGYW programs to better meet the sexual and reproductive health needs of AGYW at high risk of HIV, but also investment in tailored youth-friendly strategies for AGYW who have been marginalized from the current HIV response, including young women who engage in sexual relationships with older partners and those who sell sex.

## Data Availability

Data may be made available by the corresponding author upon receipt of reasonable request.

## Acknowledgements

The authors wish to thank the participants that engaged with the Sauti Project, as well as the Sauti program staff that worked diligently for many years to deliver critical health services for key and priority populations across Tanzania.

## Supporting Information

S1 File. SAUTI Adolescent Girls and Young Women Vulnerability Index

S1 Table. Characteristics of 65,185 AGYW accessing combination HIV prevention services in Tanzania through the Sauti Project, 2015-2019

S2 Table. Unadjusted and adjusted associations of A) transactional sex with HIV prevalence; B) age-disparate sex with HIV prevalence; and C) intergenerational sex with HIV prevalence among 4,272 sexually active adolescent girls ages 15-19 accessing combination HIV services in Tanzania through the Sauti Project, 2015-2019

## Notes

### Competing Interest Statement

The authors have declared no competing interest.

### Funding Statement

KR was supported by an award from the National Institute of Mental Health (K01MH129226). KA was supported by two awards from the National Institute of Mental Health (R25MH083620, F31MH124583). SB was supported by an award from the National Institute of Allergy and Infectious Diseases (R01AI170249). SS was also supported by an award from the National Institute of Allergy and Infectious Diseases (P30AI094189, Sub-Project ID 9272).

### Author Declarations

Ethical approval for the use of de-identified routine data was provided by the National Institute for Medical Research (NIMR/HQ/R.8c/Vol.1/678) and the Johns Hopkins Institutional Review Board (IRB No. 00006673).

## References

1. Joint United Nations Programme on HIV/AIDS. UNAIDS Data 2021 [Internet]. Geneva, Switzerland; [cited 2022 Jan 4]. Available from: https://www.unaids.org/en/resources/documents/2021/2021_unaids_data

2. The DREAMS core package of interventions: A comprehensive approach to preventing HIV among adolescent girls and young women [Internet]. [cited 2020 Jul 6]. Available from: https://journals.plos.org/plosone/article?id=10.1371/journal.pone.0208167

3. Adolescents-HIV-Eastern-Southern-Africa-2021.pdf [Internet]. [cited 2022 Jun 16]. Available from: https://www.unicef.org/esa/media/8791/file/Adolescents-HIV-Eastern-Southern-Africa-2021.pdf

4. PHIA Project [Internet]. [cited 2022 Jan 5]. Tanzania. Available from: https://phia.icap.columbia.edu/countries/tanzania/

5. Brown K. Status of HIV Epidemic Control Among Adolescent Girls and Young Women Aged 15–24 Years — Seven African Countries, 2015–2017. MMWR Morb Mortal Wkly Rep [Internet]. 2018 [cited 2020 Jul 5];67. Available from: https://www.cdc.gov/mmwr/volumes/67/wr/mm6701a6.htm

6. Harrison A, Colvin CJ, Kuo C, Swartz A, Lurie M. Sustained High HIV Incidence in Young Women in Southern Africa: Social, Behavioral, and Structural Factors and Emerging Intervention Approaches. Curr HIV/AIDS Rep. 2015 Jun;12(2):207–15.

7. Comins CA, Rucinski KB, Baral S, Abebe SA, Mulu A, Schwartz SR. Vulnerability profiles and prevalence of HIV and other sexually transmitted infections among adolescent girls and young women in Ethiopia: A latent class analysis. PloS One. 2020;15(5):e0232598.

8. Delany-Moretlwe S, Cowan FM, Busza J, Bolton-Moore C, Kelley K, Fairlie L. Providing comprehensive health services for young key populations: needs, barriers and gaps. J Int AIDS Soc. 2015;18(2 Suppl 1):19833.

9. Pettifor AE, Measham DM, Rees HV, Padian NS. Sexual power and HIV risk, South Africa. Emerg Infect Dis. 2004;10(11):1996.

10. Brook DW, Morojele NK, Zhang C, Brook JS. South African adolescents: pathways to risky sexual behavior. AIDS Educ Prev. 2006;18(3):259–72.

11. Sanders RA. Adolescent psychosocial, social, and cognitive development. Pediatr Rev. 2013 Aug;34(8):354–8; quiz 358–9.

12. Jewkes RK, Dunkle K, Nduna M, Shai N. Intimate partner violence, relationship power inequity, and incidence of HIV infection in young women in South Africa: a cohort study. The Lancet. 2010 Jul 3;376(9734):41–8.

13. Harrison A, O’Sullivan LF, Hoffman S, Dolezal C, Morrell R. Gender role and relationship norms among young adults in South Africa: measuring the context of masculinity and HIV risk. J Urban Health Bull N Y Acad Med. 2006 Jul;83(4):709–22.

14. Gottert A, Barrington C, McNaughton-Reyes HL, Maman S, MacPhail C, Lippman SA, et al. Gender Norms, Gender Role Conflict/Stress and HIV Risk Behaviors Among Men in Mpumalanga, South Africa. AIDS Behav. 2018 Jun 1;22(6):1858–69.

15. Palermo T, Chzhen Y, Balvin N, Kajula L, Palermo T, Groppo V, et al. Examining determinants of gender attitudes: evidence among Tanzanian adolescents. BMC Womens Health. 2020 Sep 10;20(1):195.

16. Rucinski KB, Schwartz SR, Mishra S, Phaswana-Mafuya N, Diouf D, Mothopeng T, et al. High HIV Prevalence and Low HIV-Service Engagement Among Young Women Who Sell Sex: A Pooled Analysis Across 9 Sub-Saharan African Countries. J Acquir Immune Defic Syndr 1999. 2020 Oct 1;85(2):148–55.

17. Hokororo A, Kihunrwa A, Hoekstra P, Kalluvya SE, Changalucha JM, Fitzgerald DW, et al. High prevalence of sexually transmitted infections in pregnant adolescent girls in Tanzania: a multi-community cross-sectional study. Sex Transm Infect. 2015 Nov;91(7):473–8.

18. Erulkar A, Medhin G, Weissman E. The impact and cost of child marriage prevention in rural Tanzania. Poverty Gend Youth [Internet]. 2017 Jan 1; Available from: https://knowledgecommons.popcouncil.org/departments_sbsr-pgy/536

19. Topazian HM, Stoner MCD, Edwards JK, Kahn K, Gómez-Olivé FX, Twine R, et al. Variations in HIV Risk by Young Women’s Age and Partner Age Disparity in Rural South Africa (HPTN 068). J Acquir Immune Defic Syndr 1999. 2020 Apr 1;83(4):350–6.

20. de Oliveira T, Kharsany ABM, Gräf T, Cawood C, Khanyile D, Grobler A, et al. Transmission networks and risk of HIV infection in KwaZulu-Natal, South Africa: a community-wide phylogenetic study. Lancet HIV. 2017 Jan;4(1):e41–50.

21. Okiring J, Getahun M, Gutin SA, Lebu S, Lee J, Maeri I, et al. Sexual partnership concurrency and age disparities associated with sexually transmitted infection and risk behavior in rural communities in Kenya and Uganda. Int J Infect Dis. 2022 Jul 1;120:158– 67.

22. McCloskey LA, Eloff I, Doran K. Determinants of intergenerational sexual relationships and HIV risk among South African women outpatients in Gauteng. AIDS Care. 2021 May;33(5):654–62.

23. Kilburn K, Ranganathan M, Stoner MCD, Hughes JP, MacPhail C, Agyei Y, et al. Transactional sex and incident HIV infection in a cohort of young women from rural South Africa. AIDS Lond Engl. 2018;32(12):1669–77.

24. Jewkes R, Dunkle K, Nduna M, Shai NJ. Transactional sex and HIV incidence in a cohort of young women in the stepping stones trial. J AIDS Clin Res. 2012;3(5).

25. Wamoyi J, Stobeanau K, Bobrova N, Abramsky T, Watts C. Transactional sex and risk for HIV infection in sub-Saharan Africa: a systematic review and meta-analysis. J Int AIDS Soc. 2016;19(1):20992.

26. Stoebenau K, Heise L, Wamoyi J, Bobrova N. Revisiting the understanding of “transactional sex” in sub-Saharan Africa: A review and synthesis of the literature. Soc Sci Med. 2016 Nov 1;168:186–97.

27. Wamoyi J, Heise L, Meiksin R, Kyegombe N, Nyato D, Buller AM. Is transactional sex exploitative? A social norms perspective, with implications for interventions with adolescent girls and young women in Tanzania. Fielding-Miller RK, editor. PLOS ONE. 2019 Apr 2;14(4):e0214366.

28. Ranganathan M, Kilburn K, Stoner MCD, Hughes JP, MacPhail C, Gomez-Olive FX, et al. The Mediating Role of Partner Selection in the Association Between Transactional Sex and HIV Incidence Among Young Women. J Acquir Immune Defic Syndr 1999. 2020 01;83(2):103–10.

29. Stoner MCD, Nguyen N, Kilburn K, Gómez-Olivé FX, Edwards JK, Selin A, et al. Age-disparate partnerships and incident HIV infection in adolescent girls and young women in rural South Africa. AIDS Lond Engl. 2019 27;33(1):83–91.

30. Rucinski KB, Kilburn K, delong, Stephanie Marie, Gómez-Olivé FX, Kahn K, Twine R, et al. Developmental trajectories of transactional sex and age-disparate relationships during adolescence: An HPTN 068 analysis. 2019 Jul; Mexico City, Mexo.

31. Full article: Sexual health of adolescent girls and young women in Central Uganda: exploring perceived coercive aspects of transactional sex [Internet]. [cited 2022 Jul 15]. Available from: https://www.tandfonline.com/doi/full/10.1080/26410397.2019.1700770

32. Becker ML, Bhattacharjee P, Blanchard JF, Cheuk E, Isac S, Musyoki HK, et al. Vulnerabilities at First Sex and Their Association With Lifetime Gender-Based Violence and HIV Prevalence Among Adolescent Girls and Young Women Engaged in Sex Work, Transactional Sex, and Casual Sex in Kenya. J Acquir Immune Defic Syndr 1999. 2018 01;79(3):296–304.

33. Wamoyi J, Wight D, Plummer M, Mshana GH, Ross D. Transactional sex amongst young people in rural northern Tanzania: an ethnography of young women’s motivations and negotiation. Reprod Health. 2010 Apr 29;7(1):2.

34. Gichane MW, Wamoyi J, Atkins K, Balvanz P, Maman S, Majani E, et al. The influence of cash transfers on engagement in transactional sex and partner choice among adolescent girls and young women in Northwest Tanzania. Cult Health Sex. 2020 Sep 16;1–15.

35. Mbita G, Mwanamsangu A, Plotkin M, Casalini C, Shao A, Lija G, et al. Consistent Condom Use and Dual Protection Among Female Sex Workers: Surveillance Findings from a Large-Scale, Community-Based Combination HIV Prevention Program in Tanzania. AIDS Behav. 2019 Aug 23;

36. Mbita G, Komba AN, Casalini C, Bazant E, Curran K, Christensen A, et al. Predictors of HIV Among 1 Million Clients in High-Risk Male Populations in Tanzania. AIDS Behav. 2022 Apr 1;1–14.

37. Nyato D, Nnko S, Komba A, Kuringe E, Plotkin M, Mbita G, et al. Facilitators and barriers to linkage to HIV care and treatment among female sex workers in a community-based HIV prevention intervention in Tanzania: A qualitative study. PLOS ONE. 2019 Nov 19;14(11):e0219032.

38. Kuringe E, Materu J, Nyato D, Majani E, Ngeni F, Shao A, et al. Prevalence and correlates of depression and anxiety symptoms among out-of-school adolescent girls and young women in Tanzania: A cross-sectional study. PLOS ONE. 2019 Aug 16;14(8):e0221053.

39. Wambura M, Drake M, Kuringe E, Majani E, Nyato D, Casalini C, et al. Cash Transfer to Adolescent Girls and Young Women to Reduce Sexual Risk Behavior (CARE): Protocol for a Cluster Randomized Controlled Trial. JMIR Res Protoc. 2019 Dec 20;8(12):e14696.

40. Han H, Yang F, Murray S, Mbita G, Bangser M, Rucinski K, et al. Characterizing a sexual health and HIV risk stratification scale for sexually active adolescent girls and young women (AGYW) in Tanzania. PloS One. 2021;16(3):e0248153.

41. Stoebenau K, Heise L, Wamoyi J, Bobrova N. Revisiting the understanding of “transactional sex” in sub-Saharan Africa: A review and synthesis of the literature. Soc Sci Med. 2016 Nov;168:186–97.

42. Leclerc-Madlala S. Age-disparate and intergenerational sex in southern Africa: the dynamics of hypervulnerability. AIDS Lond Engl. 2008 Dec;22 Suppl 4:S17–25.

43. UNAIDS Terminology Guidelines - 2015 [Internet]. [cited 2023 Jan 9]. Available from: https://www.unaids.org/en/resources/documents/2015/2015_terminology_guidelines

44. Knol MJ, VanderWeele TJ. Recommendations for presenting analyses of effect modification and interaction. Int J Epidemiol. 2012 Apr 1;41(2):514–20.

45. Howe CJ, Cole SR, Lau B, Napravnik S, Eron JJ. Selection Bias Due to Loss to Follow Up in Cohort Studies. Epidemiol Camb Mass. 2016 Jan;27(1):91–7.

46. Cole SR, Hernán MA. Constructing inverse probability weights for marginal structural models. Am J Epidemiol. 2008 Sep 15;168(6):656–64.

47. Stuart EA, Azur M, Frangakis C, Leaf P. Multiple Imputation With Large Data Sets: A Case Study of the Children’s Mental Health Initiative. Am J Epidemiol. 2009 May 1;169(9):1133– 9.

48. Azur MJ, Stuart EA, Frangakis C, Leaf PJ. Multiple imputation by chained equations: what is it and how does it work? Int J Methods Psychiatr Res. 2011 Feb 24;20(1):40–9.

49. Gichane MW, Rosenberg NE, Zimmer C, Pettifor AE, Maman S, Maseko B, et al. Individual and Relationship-Level Correlates of Transactional Sex Among Adolescent Girls and Young Women in Malawi: A Multilevel Analysis. AIDS Behav. 2022;26(3):822–32.

50. Birdthistle I, Kwaro D, Shahmanesh M, Baisley K, Khagayi S, Chimbindi N, et al. Evaluating the impact of DREAMS on HIV incidence among adolescent girls and young women: A population-based cohort study in Kenya and South Africa. PLoS Med. 2021 Oct;18(10):e1003837.

51. Iwelunmor J, Nwaozuru U, Obiezu-Umeh C, Uzoaru F, Ehiri J, Curley J, et al. Is it time to RE-AIM? A systematic review of economic empowerment as HIV prevention intervention for adolescent girls and young women in sub-Saharan Africa using the RE-AIM framework. Implement Sci Commun. 2020 Jun 10;1(1):53.

52. Groves AK, Maman S, Stankard PH, Gebrekristos LT, Amon JJ, Moodley D. Addressing the unique needs of adolescent mothers in the fight against HIV. J Int AIDS Soc. 2018 Jun 29;21(6):e25155.

53. Kasirye R, Laurenzi C, Nabulya A, Nakijoba B. Safe spaces, vocational training, and prevention programs protect young Ugandan women: findings from Uganda Youth Development Link’s DREAMS initiative for rural communities. Vulnerable Child Youth Stud. 2023 Apr 3;18(2):195–206.

54. Kangaude G, Coast E, Fetters T. Adolescent sexual and reproductive health and universal health coverage: a comparative policy and legal analysis of Ethiopia, Malawi and Zambia. Sex Reprod Health Matters. 28(2):1832291.

55. Gichane MW, Moracco KE, Pettifor AE, Zimmer C, Maman S, Phanga T, et al. Socioeconomic Predictors of Transactional Sex in a Cohort of Adolescent Girls and Young Women in Malawi: A Longitudinal Analysis. AIDS Behav. 2020 Dec 1;24(12):3376–84.

56. Busza J, Mtetwa S, Mapfumo R, Hanisch D, Wong-Gruenwald R, Cowan F. Underage and underserved: reaching young women who sell sex in Zimbabwe. AIDS Care. 2016;28 Suppl 2:14–20.

57. Napierala S, Chabata ST, Fearon E, Davey C, Hargreaves J, Busza J, et al. Engagement in HIV Care Among Young Female Sex Workers in Zimbabwe. J Acquir Immune Defic Syndr 1999. 2018 01;79(3):358–66.

58. Chabata ST, Makandwa R, Hensen B, Mushati P, Chiyaka T, Musemburi S, et al. Strategies to Identify and Reach Young Women Who Sell Sex With HIV Prevention and Care Services: Lessons Learnt From the Implementation of DREAMS Services in Two Cities in Zimbabwe. JMIR Public Health Surveill. 2022 Jul 27;8(7):e32286.

59. Chabata ST, Hensen B, Chiyaka T, Mushati P, Musemburi S, Dirawo J, et al. The impact of the DREAMS partnership on HIV incidence among young women who sell sex in two Zimbabwean cities: results of a non-randomised study. BMJ Glob Health. 2021 Apr 1;6(4):e003892.

60. Bhattacharjee P, Musau A, Manguro G, Ongwen P, Mutegi J, Kioko J, et al. HIV prevention programme with young women who sell sex in Mombasa, Kenya: learnings for scale-up. J Int AIDS Soc. 2022;25(8):e25969.

61. Hensen B, Chabata ST, Floyd S, Chiyaka T, Mushati P, Busza J, et al. HIV risk among young women who sell sex by whether they identify as sex workers: analysis of respondent-driven sampling surveys, Zimbabwe, 2017. J Int AIDS Soc [Internet]. 2019 Dec 3 [cited 2020 May 9];22(12). Available from: https://www.ncbi.nlm.nih.gov/pmc/articles/PMC6887898/

62. Ranganathan M, MacPhail C, Pettifor A, Kahn K, Khoza N, Twine R, et al. Young women’s perceptions of transactional sex and sexual agency: a qualitative study in the context of rural South Africa. BMC Public Health. 2017 Aug 22;17:666.

63. Milovanovic M, Jewkes R, Matuludi M, Dunkle K, Hlongwane K, Vanleeuw L, et al. Sex work and young women: a cross sectional study to understand the overlap of age and sex work as a central tenet to epidemic control in South Africa. AIDS Care. 2022 Apr 4;1–9.

64. Bowring AL, Ketende S, Rao A, Njindam IM, Decker MR, Lyons C, et al. Characterising unmet HIV prevention and treatment needs among young female sex workers and young men who have sex with men in Cameroon: a cross-sectional analysis. Lancet Child Adolesc Health. 2019 Jul 1;3(7):482–91.

65. Degtiar I, Rose S. A Review of Generalizability and Transportability. Annu Rev Stat Its Appl. 2023;10(1):null.

66. HIV and adolescents: focus on young key populations - Bekker - 2015 - Journal of the International AIDS Society - Wiley Online Library [Internet]. [cited 2022 Jul 15]. Available from: https://onlinelibrary.wiley.com/doi/full/10.7448/IAS.18.2.20076

67. Cheuk E, Isac S, Musyoki H, Pickles M, Bhattacharjee P, Gichangi P, et al. Informing HIV Prevention Programs for Adolescent Girls and Young Women: A Modified Approach to Programmatic Mapping and Key Population Size Estimation. JMIR Public Health Surveill. 2019 Apr 1;5(2):e11196.

68. Rucinski K, Masankha Banda L, Olawore O, Akolo C, Zakaliya A, Chilongozi D, et al. HIV Testing Approaches to Optimize Prevention and Treatment for Key and Priority Populations in Malawi. Open Forum Infect Dis. 2022 Apr 1;9(4):ofac038.

69. Olawore O, Astatke H, Lillie T, Persaud N, Lyons C, Kamali D, et al. Peer Recruitment Strategies for Female Sex Workers Not Engaged in HIV Prevention and Treatment Services in Côte d’Ivoire: Program Data Analysis. JMIR Public Health Surveill. 2020;6(4):e18000.

70. Ortblad KF, Mawandia S, Bakae O, Tau L, Grande M, Mogomotsi GP, et al. Using routine programmatic data to measure HIV incidence among pregnant women in Botswana. Popul Health Metr. 2022 Mar 4;20:10.

71. Rao A, Mhlophe H, Comins C, Young K, Mcingana M, Lesko C, et al. Persistence on oral pre-exposure prophylaxis (PrEP) among female sex workers in eThekwini, South Africa, 2016-2020. PloS One. 2022;17(3):e0265434.

72. Low A, Gummerson E, Schwitters A, Bonifacio R, Teferi M, Mutenda N, et al. Food insecurity and the risk of HIV acquisition: findings from population-based surveys in six sub-Saharan African countries (2016-2017). BMJ Open. 2022 Jul 12;12(7):e058704.

